# Predictors of Frequent Acute Respiratory Infections in Children with Tracheostomies

**DOI:** 10.64898/2026.01.28.26345051

**Authors:** Nicole Xia, Jennifer Henningfeld, Rebecca Steuart

**Affiliations:** Department of Pediatrics, Medical College of Wisconsin; Section of Pulmonary and Sleep Medicine, Department of Pediatrics, Medical College of Wisconsin; Section of Complex Care, Department of Pediatrics, Medical College of Wisconsin; Complex Care Program, Children’s Wisconsin

**Keywords:** Tracheostomy, acute respiratory infection, *Pseudomonas*

## Abstract

**Background:** Children with tracheostomies experience frequent and recurrent acute respiratory infections (ARIs). While cultured respiratory pathogens can inform ARI diagnosis, it is unknown if their presence in the airway affects future ARI risk.

**Objective:** To identify predictors of frequent (3+) ARIs within 36 months of tracheostomy.

**Methods:** We conducted a single-center, retrospective cohort study of children with tracheostomies placed between 2010-2016. Medical records were reviewed for each encounter in which a respiratory culture was obtained over the 3 years post-tracheostomy. ARIs were defined using encounter ICD-9/10 codes. Logistic and Poisson regression were used to model the association between clinical and microbiologic predictor variables with having frequent (3+) ARIs and the total number of ARIs per child. Mediation analysis using stepwise regression models further evaluated the role of *P. aeruginosa*.

**Results:** Among 436 children, 631 diagnosed ARIs occurred within 36 months of tracheostomy; 20.2% of children had 3+ ARIs. *Pseudomonas aeruginosa* was isolated in 25% of children and was more common among those with 3+ ARIs compared with 0-2 ARIs (56.8% vs 20.7%, p<0.001). Those with early *P. aeruginosa* isolation were more likely to have 3+ ARIs (aOR 3.38, 95% CI 1.97-5.81), and this relationship persisted when analyzing ARIs and *P. aeruginosa* counts. Identification of *P. aeruginosa* partially mediated the relationship of ventilator dependence with ARI frequency.

**Conclusion:** Isolation of *P. aeruginosa*, particularly early and repeated isolation, is associated with frequent ARIs in the 3 years after tracheostomy and is an important partial mediator. Findings may inform risk stratification and targeted treatment strategies.

## Background

Children with tracheostomies are at high risk for acute respiratory infection (ARI), a common cause for hospitalization, readmission, and mortality in this population.^1-5^ One presumed mechanism for ARI risk is that the tracheostomy bypasses normal upper airway defenses, facilitating direct access of pathogens to the lower respiratory tract and predisposing to both colonization and infection.^6^ The tracheostomy tube itself, and the associated local inflammatory response of the airway,^7^ may further impair mucosal defenses and thus increase infection risk. Additional ARI risk factors include underlying airway, pulmonary, or neurologic disease, as well as co-morbid chronic conditions.^2-3^

Respiratory cultures are frequently obtained in children with tracheostomies presenting with new or worsening respiratory symptoms to guide ARI diagnosis and therapy.^8-9^ However, the diagnostic utility of these cultures can be limited. During ARI, our prior work demonstrates that the sensitivity of respiratory cultures for bacterial ARI is low and the positive predictive value is modest, reflecting the challenge of distinguishing bacterial colonization of the airway from infection.^10^ Furthermore, routine surveillance cultures in asymptomatic children also poorly predict pathogens during subsequent ARIs and may contribute to unnecessary antibiotic use.^9^ Surveillance cultures, on the other hand, may be used to evaluate for bacterial colonization of the lower airways. Commonly isolated organisms include *Pseudomonas aeruginosa, Staphylococcus aureus*, and *Serratia marcescens*, with *P. aeruginosa* identified in up to 90% of respiratory cultures in some cohorts.^11-14^ In other lung diseases including cystic fibrosis, *P. aeruginosa* colonization is associated with more lung exacerbations, declining lung function, poorer quality of life, and increased health care costs related to medications and hospitalization.^15^ However, the clinical significance of these organisms, particularly *P. aeruginosa*, in children with tracheostomies and their relationship to future infection risk remains unclear. Understanding whether early or repeated isolation of *P. aeruginosa* is associated with subsequent respiratory illness could help refine monitoring and antibiotic stewardship strategies.

### Objectives

We aimed to identify clinical and microbiologic predictors associated with a child contracting frequent (i.e., 3+) ARIs within 36 months of tracheostomy placement.

## Methods

### Study Design, Population, and Data Source

This was a single center, retrospective cohort study using data from Cincinnati Children’s Hospital Medical Center (CCHMC). Children 2 months–18 years old with tracheostomies placed between January 2010-December 2016 were included, as identified by an internal tracheostomy patient registry maintained by the Division of Pulmonary Medicine. Children with cystic fibrosis were excluded due to their distinct airway microbiology. For the 36 months following tracheostomy placement, or December 2018 (whichever occurred first), detailed demographic, clinical, and respiratory culture data was extracted from the electronic medical record (EMR) for all encounters in which a respiratory culture (tracheostomy aspirate or bronchoalveolar lavage) was collected. For children with multiple respiratory cultures collected in the same encounter, the initial culture was collected. Inpatient, outpatient, and lab collection only encounters were included. Respiratory cultures could be obtained for any reason, including illness evaluation or routine surveillance.

This study was reviewed by the CCHMC and Children’s Wisconsin Institutional Review Boards and determined to be exempt research (CCHMC IRB ID 2019-0938, CW IRBNet ID 1805584), without the requirement for informed consent. This determination was based on the use of existing medical records without additional patient interventions, and that all data were de-identified to ensure patient privacy.

### Predictors and outcome measures

We evaluated demographic, clinical, and microbiological variables hypothesized to contribute to ARI susceptibility for univariate significance. The demographic variables evaluated included age at tracheostomy placement, race, ethnicity, and insurance type listed in the EMR at the time of data extraction. Microbiological predictors evaluated included the presence of *P. aeruginosa* and *S. aureus* isolation, respectively, in a respiratory culture within the first 36 months after tracheostomy placement. For primary analysis, *P. aeruginosa* and *S. aureus* organism isolations were defined dichotomously (presence/absence within the 36 months). For secondary analyses of the association of *P. aeruginosa* specifically, organism isolations were defined as a) a count of culture instances in which *P. aeruginosa* was isolated and b) presence/absence of early *P. aeruginosa*, defined as a culture isolation within 12 months of tracheostomy placement.

The primary outcome was a child having 3+ encounters in which an ARI was diagnosed and respiratory culture was collected within 36 months of the tracheostomy placement. The secondary outcome was the number of encounters with diagnosed ARIs within 36 months. ARI was defined using ICD-9 and ICD-10 diagnostic codes placed by clinicians using our group’s previously defined diagnostic coding scheme.^10^

### Covariates

Demographic variables of sex, race, ethnicity, and insurance status were measured from the EMR at the time of data collection, as was the patient age at tracheostomy placement and at decannulation or death, if applicable. Clinical variables evaluated included each child’s number of organ systems with complex chronic conditions (CCCs), CCC category, chronic ventilator dependence, bronchopulmonary dysplasia (BPD) diagnosis, and high intensity neurological impairment (HI-NI) diagnosis. BPD and HI-NI were defined using previously validated *International Classification of Diseases, Ninth Revision* (ICD-9) and *Tenth Revision* (ICD-10) codes.^16-17^ Diagnosis of BPD, HI-NI, and number and category of CCCs were collected at each encounter and the maximum value across encounters was used to define each child’s status. Chronic ventilator dependence was defined in the Pulmonary Medicine database during outpatient Pulmonary clinic visits as regular use of a ventilator at home for any portion of the day or night.

### Statistical Analysis

Chi-square and Wilcoxon rank sum tests were used to examine differences in predictor variables between primary outcome groups. In adjusted analysis, logistic regression equations were used to determine association of significant identified predictors and covariates with our primary outcome of experiencing 3+ ARI within 36 months. Poisson regression models were used to determine the association of these predictors with our secondary outcome, the count of diagnosed ARIs within 36 months. To further explore the relationship of *P. aeruginosa* identification with the primary outcome, logistic regression and Poisson regression models additionally evaluated associations of the predictor early *P. aeruginosa* with primary and secondary outcomes. Number of cultures obtained per child was not included in regression models as it was determined to be a collider variable.

A mediation analysis was conducted using Baron and Kenny’s stepwise regression model approach^18^ to examine whether a child having isolation of *P. aeruginosa* explains the relationship between chronic ventilator dependence and 3+ ARIs within 36 months. The Sobel test was used to confirm statistical significance of the mediating relationship.

As a sensitivity analysis, children who had less than 2 years of respiratory culture and encounter data were excluded; specifically, children who decannulated or died within 2 years of tracheostomy placement and children who had no respiratory cultures collected after 2 years of tracheostomy placement (potentially lost to follow-up) were excluded.

Analyses were performed with R v4.1.1 (Vienna, Austria). *P*-values <0.05 were considered statistically significant.

## Results

This study identified 436 children with tracheostomies during the 9–year study period. The cohort experienced 631 diagnosed ARIs within 3 years of tracheostomy placement. Children had a median 1 ARI per child, interquartile range (IQR): 0–2, full range: 0–11. Among these children, 88 (20.2%) had 3+ diagnosed ARIs (**Table 1**).

**Table 1.** Population characteristics.

|  | All children<br>(*n*=431) |  | Children with 0-2 ARIs<br>within 36 months<br>(*n*=348) |  | Children with 3+ ARIs<br>within 36 months<br>(*n*=88) |  |  |
| --- | --- | --- | --- | --- | --- | --- | --- |
| Risk Factor | number | percentage | number | percentage | number | percentage | p-value |
| **Demographic and clinical characteristics** |  |  |  |  |  |  |  |
| Male | 244 | 56.6% | 191 | 54.9% | 53 | 60.2% | 0.43 |
| Race |  |  |  |  |  |  | 0.67 |
| White | 295 | 68.5% | 234 | 67.2% | 61 | 69.3% |  |
| Black or AA | 95 | 22.0% | 75 | 21.6% | 20 | 22.7% |  |
| Other* | 46 | 10.7% | 39 | 11.2% | 7 | 8.0% |  |
| Hispanic | 15 | 3.5% | 9 | 2.6% | 6 | 6.8% | 0.11 |
| Insurance type** |  |  |  |  |  |  | **0.01** |
| Public | 213 | 49.4% | 161 | 46.3% | 52 | 59.1% |  |
| Private | 204 | 47.3% | 168 | 48.3% | 36 | 40.9% |  |
| Other, uninsured | 19 | 4.4% | 19 | 5.5% | 0 | 0% |  |
| Age at tracheostomy placement (median, IQR) | 5 months | (3,29) | 5 months | (3,50) | 3.5 months | (2,8) | **<0.001** |
| Chronic ventilator dependence | 210 | 48.7% | 142 | 40.8% | 68 | 77.3% | **<0.001** |
| Bronchopulmonary dysplasia (BPD) or Chronic Lung Disease of Infancy (CLDI) diagnosis | 136 | 31.6% | 110 | 31.6% | 26 | 29.6% | 0.74 |
| Higher-intensity Neurologic Impairment (HI-NI) diagnosis | 271 | 62.9% | 204 | 58.6% | 67 | 76.1% | **0.004** |
| Number of Complex Chronic Condition categories (CCCs) (median, IQR) | 4 | (3,6) | 4 | (3,6) | 5 | (4,6) | **0.01** |
| Category of CCC |  |  |  |  |  |  |  |
| Respiratory | 373 | 86.5% | 293 | 84.2% | 80 | 90.9% | 0.15 |
| Gastrointestinal | 288 | 66.8% | 217 | 62.4% | 71 | 80.7% | **0.002** |
| Neuromuscular | 160 | 37.1% | 124 | 35.6% | 36 | 40.9% | 0.43 |
| Cardiovascular | 187 | 43.4% | 144 | 41.4% | 43 | 48.9% | 0.25 |
| Genetic | 144 | 33.4% | 107 | 30.8% | 37 | 42.1% | 0.06 |
| Neonatal | 117 | 27.1% | 96 | 27.6% | 21 | 23.9% | 0.50 |
| Metabolic | 68 | 15.8% | 50 | 14.4% | 18 | 20.5% | 0.21 |
| Hematologic | 56 | 13.0% | 46 | 13.2% | 10 | 11.4% | 0.77 |
| Transplant | 39 | 9.0% | 28 | 8.1% | 11 | 12.5% | 0.27 |
| Renal | 57 | 13.2% | 40 | 11.5% | 17 | 19.3% | 0.08 |
| Malignancy | 32 | 7.4% | 26 | 7.5% | 6 | 6.8% | 0.99 |
| Decannulated within 36 months | 18 | 4.2% | 15 | 4.3% | 3 | 3.4% | 0.80 |
| Deceased within 36 months | 78 | 18.1% | 69 | 19.8% | 9 | 10.2% | **0.005** |
| **Respiratory culture results** |  |  |  |  |  |  |  |
| Number of cultures taken within 36 months (median, IQR) | 4 | (1,8) | 3 | (1,5) | 11 | (7,13.25) | **<0.001** |
| Any *P. aeruginosa* | 108 | 25.1% | 72 | 20.7% | 50 | 56.8% | **<0.001** |
| Count of *P. aeruginosa* (median, IQR) | 0 | (0,0.5) | 0 | (0,0) | 1 | (0,2) | **0.002** |
| Any early*** *P. aeruginosa* | 78 | 18.1% | 44 | 12.6% | 34 | 38.6% | **<0.001** |
| Any *S. aureus* | 76 | 17.6% | 51 | 14.7% | 25 | 28.4% | **0.004** |
| Any MRSA | 27 | 6.3% | 17 | 4.9% | 10 | 11.4% | **0.045** |

Abbreviations: AA: African American, ARI: Acute respiratory infection, IQR: interquartile range, MRSA: methicillin-resistant *S. aureus*.
\*Other race includes: 15 children of race “Other”, 15 of unknown race, 7 identified as Asian, 2 identified as American Indian and Alaska Native, 2 identified as Native Hawaiian and Other Pacific Islander, and 1 refused.
\*\*If multiple types of insurance (ie, dual insurance), private was selected.
\*\*\**P. aeruginosa* isolation on tracheostomy culture taken within 12 months of tracheostomy placement.

In our cohort, children were 56.6% male, with a race distribution of 68.5% White, 22.0% Black or African American, and 10.7% identified as Other (which included more than one racial category, Unknown, and Refused, **Table 1**). Half (49.4%) of children were primarily publicly-insured. There were no statistically significant differences between race, ethnicity, or gender demographic covariates between groups in our study, except having public insurance was more frequent among children with 3+ ARIs (p=0.01). Children often had comorbid conditions, with 31.6% of children having BPD and 62.9% having a HI-NI. Almost half of the children used a chronic ventilator (48.7%).

Children with 3+ ARIs versus 0-2 ARIs within 36 months were significantly more likely to have chronic ventilator dependence (76.9% vs. 43.2%, p<0.001), be younger age at tracheostomy placement (3.5 months vs 5.0 months, p<0.001), and have a HI-NI diagnosis (76.1% vs. 58.6%, p=0.004, **Table 1**). Children with 3+ ARIs also had more organ systems with CCC diagnoses (median 5 vs. 4, p<0.0001). Children with 3+ ARIs were also more likely to have a gastrointestinal CCC than children with 0-2 ARIs (80.7% vs. 62.4%, p=0.02), but there were no statistically significant differences in other categories of CCC between groups.

We identified 2,292 respiratory cultures obtained from the cohort, 14% of which were positive for bacterial organism identification and 31% which were obtained during an encounter with an ARI diagnosis. Respiratory cultures identified *P. aeruginosa* during 10% of ARIs, *S. aureus* during 6% of ARIs, and both *P. aeruginosa* and *S. aureus* during 1% of ARIs.

One quarter of children (25%, 108) had 1+ isolation of *P. aeruginosa*, most of these (78 children) having early isolation of *P. aeruginosa*. The median time to first *P. aeruginosa* isolation was 12 months (IQR 4-32 months). Among children who had 3+ ARIs, *P. aeruginosa* isolation was more frequently observed as compared with children who had 0-2 ARIs [56.8% vs. 20.7%, p<0.001, **Table 1**] including early *P. aeruginosa* isolation [38.6% vs. 12.6%, p<0.001]. *S. aureus* isolation was also more frequently observed among children with 3+ ARIs as compared with those with 0-2 ARIs [27.4% vs. 14.4%, p<0.001].

In adjusted analysis of the primary outcome, children with any *P. aeruginosa* isolation demonstrated increased odds of 3+ ARI diagnoses [adjusted odds ratio (aOR) 4.02, 95% confidence interval (CI) 2.26-7.25, **Table 2**]. The covariates of chronic ventilation use [aOR 3.09, 95% CI 1.69-5.85], HI-NI diagnosis [aOR 2.01, 95% CI 1.05-4.01], and younger age at tracheostomy placement [aOR 0.89, 95% CI 0.80-0.96] were associated with increased odds of 3+ ARIs. Additionally, those with early *P. aeruginosa* also had higher odds of 3+ ARIs [aOR 3.09, 95% CI 1.68-5.67]. Stratified analyses showed consistent effect estimates for the two levels of *P. aeruginosa* identification (presence/absence), indicating no effect modification by this variable. Any isolation of *S. aureus* post tracheostomy was not associated with 3+ ARIs compared to having 0-2 ARIs within 36 months [aOR 1.44, 95% CI 0.95-2.21].

**Table 2.**
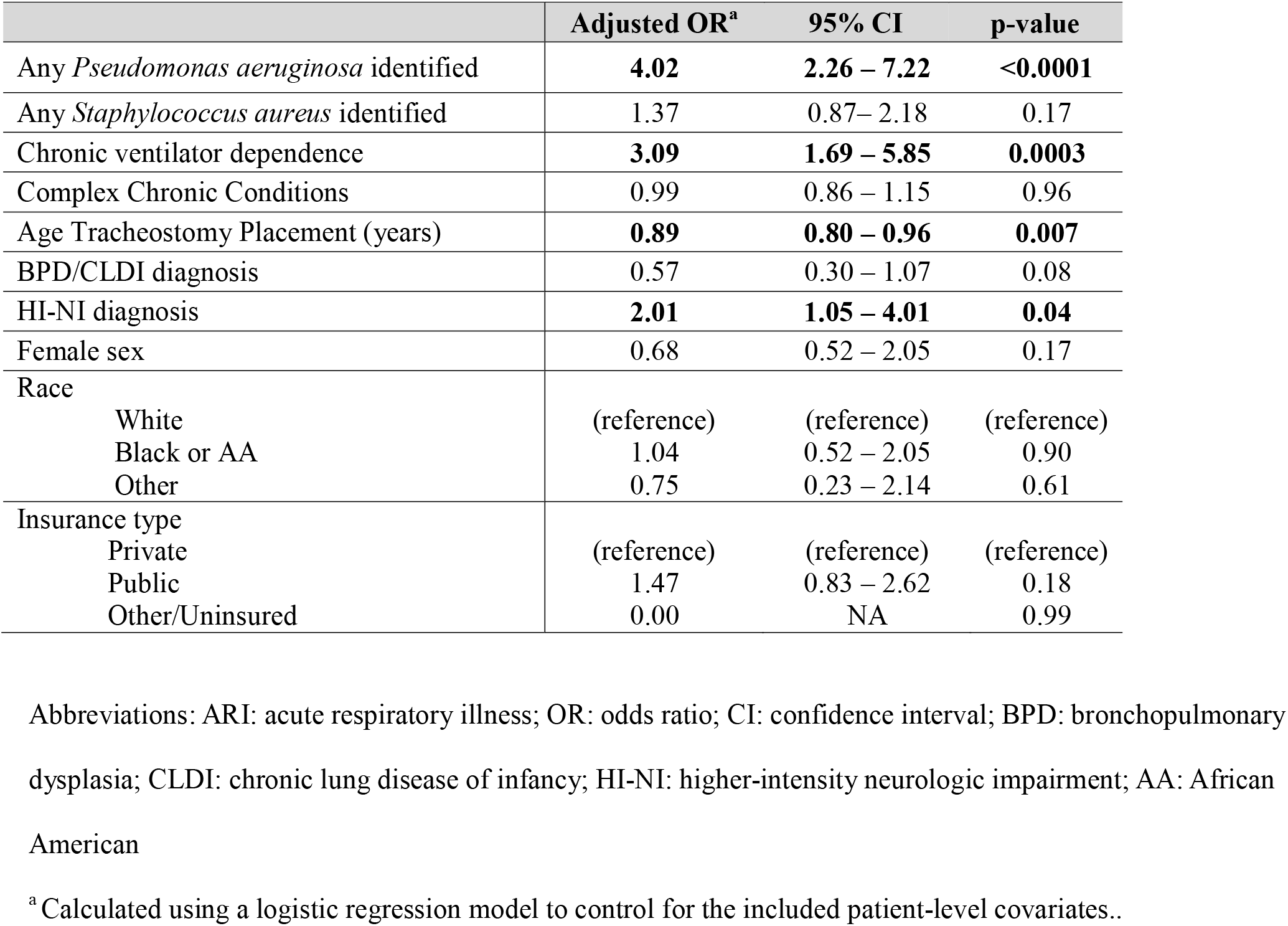
Adjusted Analysis: Characteristics associated with 3+ ARIs in the 36 months after tracheostomy placement.

|  | **Adjusted OR<sup>a</sup>** | **95% CI** | **p-value** |
| --- | --- | --- | --- |
| Any *Pseudomonas aeruginosa* identified | **4.02** | **2.26 – 7.22** | **<0.0001** |
| Any *Staphylococcus aureus* identified | 1.37 | 0.87– 2.18 | 0.17 |
| Chronic ventilator dependence | **3.09** | **1.69 – 5.85** | **0.0003** |
| Complex Chronic Conditions | 0.99 | 0.86 – 1.15 | 0.96 |
| Age Tracheostomy Placement (years) | **0.89** | **0.80 – 0.96** | **0.007** |
| BPD/CLDI diagnosis | 0.57 | 0.30 – 1.07 | 0.08 |
| HI-NI diagnosis | **2.01** | **1.05 – 4.01** | **0.04** |
| Female sex | 0.68 | 0.52 – 2.05 | 0.17 |
| Race |  |  |  |
| White | (reference) | (reference) | (reference) |
| Black or AA | 1.04 | 0.52 – 2.05 | 0.90 |
| Other | 0.75 | 0.23 – 2.14 | 0.61 |
| Insurance type |  |  |  |
| Private | (reference) | (reference) | (reference) |
| Public | 1.47 | 0.83 – 2.62 | 0.18 |
| Other/Uninsured | 0.00 | NA | 0.99 |

Abbreviations: ARI: acute respiratory illness; OR: odds ratio; CI: confidence interval; BPD: bronchopulmonary dysplasia; CLDI: chronic lung disease of infancy; HI-NI: higher-intensity neurologic impairment; AA: African American
<sup>a</sup> Calculated using a logistic regression model to control for the included patient-level covariates..

In analysis of secondary outcomes, each isolation of *P. aeruginosa* was associated with a two-fold increased odds of having frequent ARIs [adjusted risk ratio (aRR) 2.1, 95% CI 1.51-2.99, **Figure 1**]. For each early isolation of *P. aeruginosa*, there was also an increased odds of frequent ARIs [aRR of 1.27, 95% CI 1.18-1.36].

**Figure 1.**
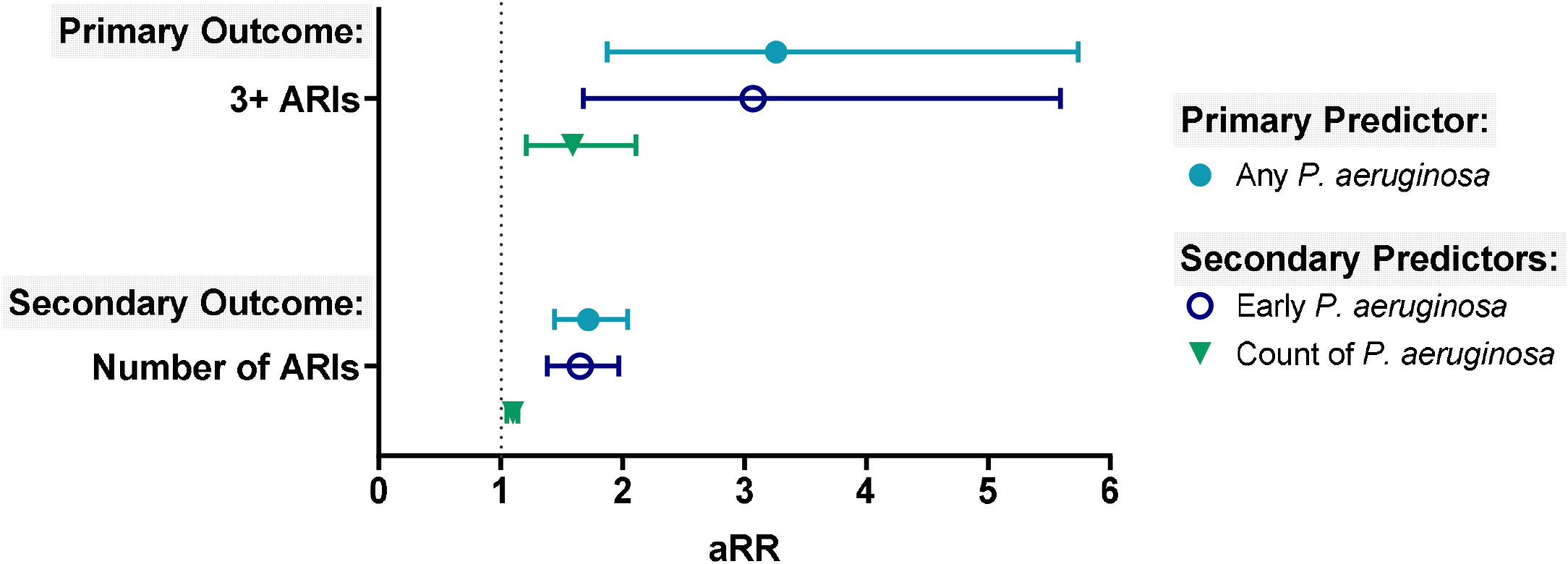
Adjusted Analyses of primary and secondary predictors and primary and secondary outcomes. Abbreviations: ARIs: Acute respiratory infections, aRR: adjusted Risk Ratio Calculated using logistic regression (primary outcome: 3+ ARIs) and Poisson regression (secondary outcome: number of ARIs), while controlling for sex, age at tracheostomy, race, insurance type, isolation of *S. aureus*, chronic ventilator dependence, number of complex chronic conditions, bronchopulmonary dysplasia diagnosis, and higher-intensity neurologic impairment diagnosis.

In mediation analysis, the indirect effect of the strongest predictor variable, chronic ventilator dependence, on the primary outcome through presence of *P. aeruginosa* isolation was significant, indicating mediation (β = 2.60, Sobel Test p-value = 0.0005, **Figure 2**). The direct effect remained significant, suggesting that *P. aeruginosa* isolation partially mediates the relationship of chronic ventilator dependence and 3+ ARIs.

**Figure 2.**
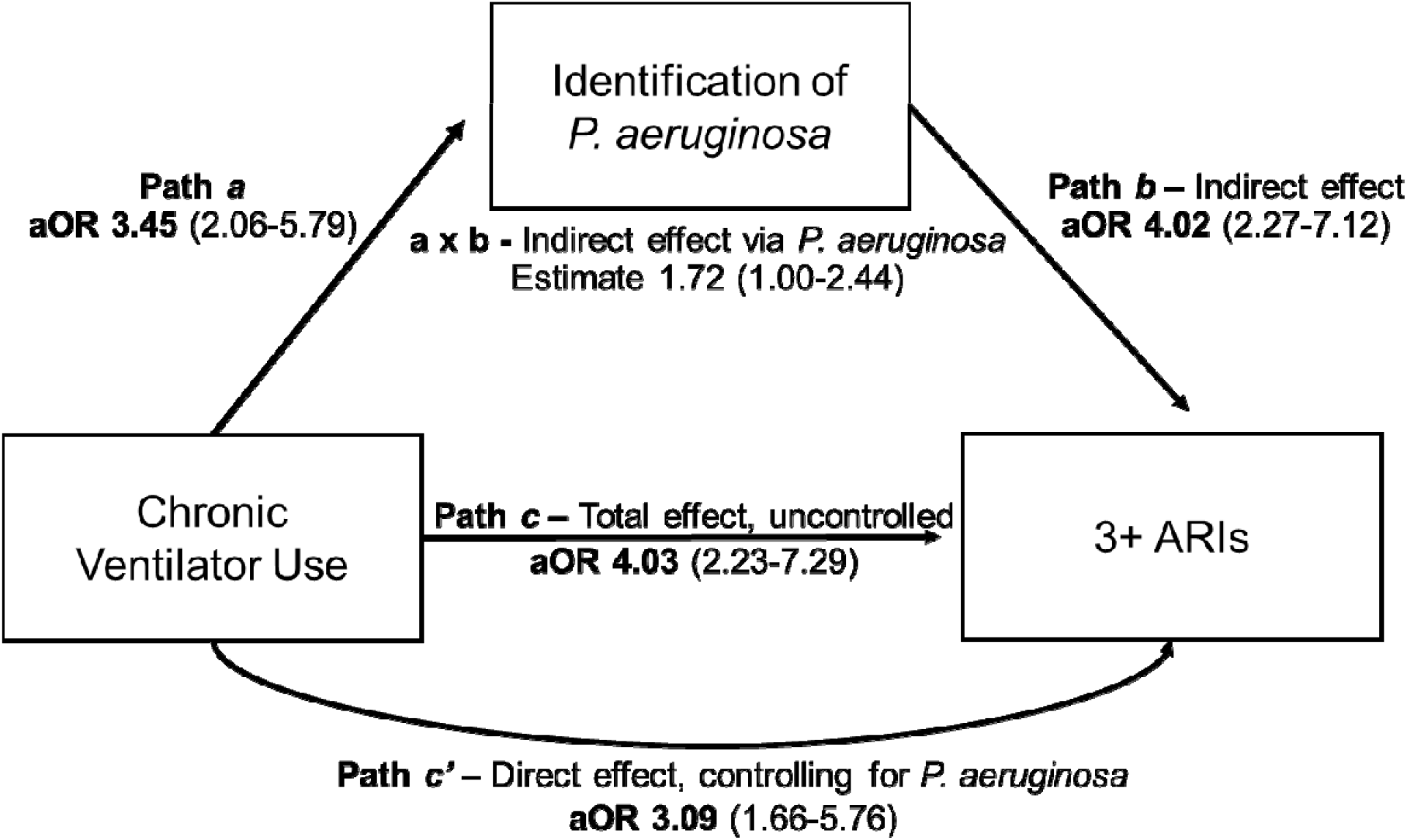
Mediation Analysis. Abbreviations: ARI: acute respiratory infection. All paths are statistically significant with p<0.001. **Interpretation:** There is a meaningful and significant mediated pathway through the variable of presence/absence of *P. aeruginosa* identification. This means part of the effect of chronic ventilator dependence on frequent (3+) ARIs is explained by its influence on *P. aeruginosa* identification. *P. aeruginosa* is a significant mediator, suggesting that interventions targeting *P. aeruginosa* may reduce ARI frequency in children with chronic ventilator dependence. ^a^ Calculated using logistic regression models, each controlling for chronic ventilator dependence, sex, age at tracheostomy, race, insurance type, isolation of *S. aureus*, number of complex chronic conditions, bronchopulmonary dysplasia diagnosis, and higher-intensity neurologic impairment diagnosis. Covariate effects values are listed in the Supplemental Table.

In the cohort, 153 children had less than 2 years of respiratory culture and encounter diagnosis data: 18 children were decannulated, 78 children died, and 108 children had no respiratory culture encounters after 2 years post-tracheostomy. In sensitivity analysis excluding these children, children with *P. aeruginosa* isolation continued to demonstrate increased odds of 3+ ARI diagnoses [aOR 3.67, 95% CI 1.89-7.25, **Table 3**]. Age of tracheostomy placement continued to be associated with 3+ ARIs [aOR 0.75, 95% CI 0.59-0.92].

**Table 3:** Sensitivity analysis: Excluding children with <2 years of data.

|  | Adjusted OR<sup>a</sup> | 95% CI | p-value |
| --- | --- | --- | --- |
| Any *Pseudomonas aeruginosa* identified | **3.67** | **1.89-7.25** | **0.0001** |
| Any *Staphylococcus aureus* identified | 1.61 | 0.97-2.71 | 0.07 |
| Chronic ventilator dependence | 1.64 | 0.79-3.49 | 0.19 |
| Complex Chronic Conditions | 1.02 | 0.86-1.21 | 0.79 |
| Age Tracheostomy Placement (years) | **0.75** | **0.59-0.92** | **0.008** |
| BPD/CLDI diagnosis | 0.61 | 0.28-1.25 | 0.18 |
| HI-NI diagnosis | 2.00 | 0.92-4.43 | 0.08 |
| Female sex | 0.55 | 0.29-1.02 | 0.06 |
| Race |  |  |  |
| White | (reference) | (reference) | (reference) |
| Black or AA | 1.27 | 0.59-2.68 | 0.54 |
| Other | 1.39 | 0.35-4.84 | 0.61 |
| Hispanic ethnicity | 3.58 | 0.82-16.40 | 0.09 |
| Insurance type |  |  |  |
| Private | (reference) | (reference) | (reference) |
| Public | 1.76 | 0.93-3.36 | 0.08 |
| Other/Uninsured | 0.00 | NA | 0.99 |

Abbreviations: ARI: acute respiratory infection; OR: odds ratio; CI: confidence interval; BPD: bronchopulmonary dysplasia; CLDI: chronic lung disease of infancy; HI-NI: higher-intensity neurologic impairment; AA: African American
Sensitivity analysis cohort included 278 children, including 203 children with 0-2 ARIs and 75 children with 3+ ARIs; 77 children had *Pseudomonas* identified, 196 without *Pseudomonas*.

## Discussion

In this retrospective cohort study, children with tracheostomies who had one or more instances of *P. aeruginosa* isolation in their respiratory cultures had higher odds of experiencing frequent pseudomonal and non-pseudomonal ARIs in the first 3 years following tracheostomy placement, even after controlling for additional ARI risk factors. This association was strongest for children with early or repeated isolations of *P. aeruginosa*, highlighting importance of both the timing and cumulative burden of colonization. Furthermore, *P. aeruginosa* acts as partial mediator for the relationship between chronic ventilator dependence and frequent ARIs, suggesting that ventilator use may facilitate colonization, which in turn contributes to infection risk. Additional factors associated with frequent ARIs included young age at tracheostomy placement and HI-NI diagnosis. In contrast, *S. aureus* isolation was not independently associated with frequent future ARIs. Our findings suggest that children with respiratory tract *P. aeruginosa* may warrant closer monitoring or targeted intervention, particularly among those with other risk factors such as chronic ventilator dependence.

*P. aeruginosa* is the most commonly isolated organism in tracheostomy aspirates of children, with reported prevalence ranging from 18-90% depending on the center.^11-14^ In this cohort, which comes from a relatively low-prevalence center, 25% of children had at least one isolation, most occurring early after tracheostomy placement. Although we identified an association of *P. aeruginosa* identification with frequent ARIs, it is noteworthy that most of the ARIs in our dataset were non-pseudomonal, regardless of prior *P. aeruginosa* identification. Furthermore, 74.1% of *P. aeruginosa* identifications occurred during wellness, consistent with our prior analysis in which *P. aeruginosa* was equally likely to be isolated during illness or wellness.^10^ Taken together, these findings suggest that *P. aeruginosa* is a colonizing organism that is not directly causing all frequent ARIs, and highlighting the need to interpret positive cultures carefully in the context of patient risk factors and clinical presentation.

In addition to evaluating dichotomous presence/absence of *P. aeruginosa*, our study examined the timing and frequency of isolation events. Early isolation of *P. aeruginosa*, defined as within the first year post-tracheostomy, was associated with higher odds of frequent ARIs compared to later isolation. This suggests that the timing of colonization is clinically meaningful. Possible mechanisms for this include longer exposure time to the effects of *P. aeruginosa*, higher *P. aeruginosa* organism burden, or higher baseline respiratory vulnerability to colonization which also confers infection risk. Our study also identified a dose-response relationship: each isolation of *P. aeruginosa* increased the odds of a child having frequent ARIs, suggesting that colonization may further increase vulnerability.

Emerging evidence indicates that colonizing respiratory bacteria have a role in maintaining respiratory health and associate with respiratory rehospitalization.^19-20^ *P. aeruginosa* frequently persists or recurs in tracheostomy aspirates despite targeted antibiotic treatment,^14^ and may sometimes be acquired prior to tracheostomy placement.^1^ The persistence of *P. aeruginosa* across wellness and illness states, and its recurrences during wellness without overt symptoms, supports its role as a colonizing airway organism.^5,20^ In other chronic lung diseases such as cystic fibrosis and chronic obstructive pulmonary disease (COPD), *P. aeruginosa* colonization is associated with increased pulmonary exacerbations.^15,21-22^ The mechanisms of these associations are less clear, however, and have been proposed to include biofilms, respiratory microbiome dysbiosis, and bacterial virulence factors.^17^ Our findings suggest a similar pattern: *P. aeruginosa* may confer vulnerability to both pseudomonal and non-pseudomonal ARIs among children with tracheostomies. However, interestingly, our study did not find *S. aureus* identification to be associated with frequent future ARIs, suggesting that not all organisms confer the same risk. Further study is needed to investigate if and how *P. aeruginosa* may play a role in decreasing respiratory tract immunity, mediating respiratory tract inflammation, and modulating respiratory microbiome dysbiosis.

Alternatively, our findings here suggest that *P. aeruginosa* identification is a marker of a vulnerable airway reflecting underlying illness severity, rather than a causative factor itself conferring vulnerability to respiratory infections. It could be that unaccounted for clinical conditions and medical complexity are themselves associated with *P. aeruginosa* identification and colonization, meaning that *P. aeruginosa* is a marker of ARI risk rather than a direct causative factor. However, the persistent association of *P. aeruginosa* with frequent ARIs in our adjusted analysis suggests that *P. aeruginosa* may have an important, independent role in respiratory bacterial systems warranting further study. Further causal inference analysis is needed to isolate the effect of respiratory bacteriology in the broader respiratory infection-colonization-technology system.

In addition to microbial factors, we confirmed three clinical risk factors – severe neurologic impairment, chronic ventilator dependence, and young age at tracheostomy – are associated with increased odds for frequent future ARIs.^2-3^ Among children with neurologic impairment, studies have shown that those with *P. aeruginosa* colonization are at higher risk for respiratory illnesses such as aspiration pneumonia and pulmonary exacerbation.^22-23^ The decreased mobility, altered upper airway clearance mechanisms, exposure to medications like proton pump inhibitors, and use of enteral feeding tubes that are common among this group are all factors thought to alter the resident microbiome,^14^ facilitating the colonization of pathogens and potential subsequent ARI development. Acute ventilator use is a known risk factor for ventilator-associated pneumonia (VAP), a prevalent nosocomial respiratory infection in pediatric critical care patients with artificial airways.^24^ Despite incomplete understanding of the pathogenesis of VAP, our study’s observation of increased ARI odds among children using chronic ventilatory support via tracheostomy aligns with this literature. It is also possible that the convergence of multiple factors together contributes to the high frequency of ARIs seen in certain children within the tracheostomy population. Closer observation and precautions for these children to monitor for early signs of acute respiratory illness could be critical in early intervention once *P. aeruginosa* is identified on respiratory culture.

The study is subject to several limitations regarding the definition and classification of ARI, as well as the accuracy of culture results. First, diagnostic codes used to define ARIs may lack sensitivity, as they are dependent on clinician-placed diagnoses, and could result in a misclassification bias of the outcome if true ARIs were not included. Our definition of ARI also included only instances in which a tracheostomy aspirate culture was obtained, potentially leading to an exposure bias. Both biases introduced by our ARI definition would be expected to bias results towards the null. Second, the primary definition of *P. aeruginosa* exposure used, which includes identification at any point in the 3 years after tracheostomy placement rather than only before the ARI outcomes, introduces an exposure bias that limits the establishment of a clear cause-and-effect relationship. This bias is mitigated in part by the identified relationship of early *P. aeruginosa* identification with ARIs. Third, the number of cultures conducted per patient differed significantly between groups; this likely introduces some bias in predictor identification, specifically, higher culture frequency could disproportionately influence the detection of certain pathogens such as *P. aeruginosa*, thus skewing the association between pathogen isolation and frequent ARIs. Additionally, variations in culture practices and thresholds for conducting cultures across different healthcare settings may impact the likelihood of identifying specific pathogens, potentially confounding the relationship between culture results and ARI frequency. Fourth, residual confounding is also a concern, as unmeasured clinical or demographic factors influencing culture acquisition and interpretation may confound the observed associations. Lastly, the study’s findings may not be generalizable beyond the study institution, given variations in culture collection practices, positive culture prevalence, and ARI diagnosis and treatment differences among different healthcare settings.

## Conclusion

*Pseudomonas aeruginosa* isolation in the respiratory tract of children with tracheostomies, particularly frequent or early isolations, is independently associated with frequent ARIs. *P. aeruginosa* partially mediates the relationship of chronic ventilator dependence, an important ARI risk factor, with ARI frequency. Our results emphasize the clinical significance of *P. aeruginosa* identification in tracheostomy aspirates, and supports its use in strategies for early detection and risk stratification to guide clinical management in this vulnerable patient population.

## Data Availability

All data produced in the present study are available upon reasonable request to the authors.

## Abbreviations

ARI: Acute Respiratory Illness
EMR: Electronic Medical Record
CCC: Complex Chronic Conditions
BPD: Bronchopulmonary Dysplasia
HI-NI: High Intensity Neurological Impairment
aOR: Adjusted Odds Ratio
CI: Confidence Interval
aRR: Adjusted Risk Ratio
COPD: Chronic Obstructive Pulmonary Disease
VAP: Ventilator-Associated Pneumonia
CLDI: Chronic Lung Disease of Infancy

---

**Supplemental Table 1.** Covariate effects across models.

**Supplemental Table. Covariate effects across models**
| Covariate | Model Path | Estimate | OR | 95% CI | p-value |
| --- | --- | --- | --- | --- | --- |
| Chronic ventilator use | a | 1.2386 | 3.45 | [2.06, 5.79] | <0.001 |
| Age at tracheostomy placement (years) | a | -0.0679 | 0.93 | [0.87, 1.00] | 0.044 |
| BPD diagnosis | a | -0.1027 | 0.90 | [0.52, 1.55] | 0.711 |
| HI-NI diagnosis | a | -0.1602 | 0.85 | [0.48, 1.50] | 0.586 |
| Male sex | a | -0.3032 | 0.74 | [0.47, 1.16] | 0.203 |
| Race (Other) | a | 0.0287 | 1.03 | [0.40, 2.67] | 0.953 |
| Race (White) | a | 0.2117 | 1.24 | [0.67, 2.29] | 0.491 |
| Non-Hispanic ethnicity | a | 0.2846 | 1.33 | [0.32, 5.52] | 0.697 |
| Insurance type (Private) | a | -1.3450 | 0.26 | [0.07, 0.91] | 0.039 |
| Insurance type (Public) | a | -1.0249 | 0.36 | [0.10, 1.25] | 0.114 |
| *S. aureus* isolation | b | 0.3167 | 1.37 | [0.87, 2.15] | 0.174 |
| Chronic ventilator use | b | 1.1283 | 3.09 | [1.66, 5.76] | <0.001 |
| Age at tracheostomy placement (years) | b | -0.1211 | 0.89 | [0.82, 0.97] | 0.007 |
| BPD diagnosis | b | -0.5607 | 0.57 | [0.30, 1.08] | 0.083 |
| HI-NI diagnosis | b | 0.6991 | 2.01 | [1.03, 3.91] | 0.042 |
| Male sex | b | 0.3878 | 1.47 | [0.85, 2.55] | 0.166 |
| Race (Other) | b | -0.3261 | 0.72 | [0.22, 2.34] | 0.587 |
| Race (White) | b | -0.0430 | 0.96 | [0.48, 1.91] | 0.902 |
| Non-Hispanic ethnicity | b | -1.4643 | 0.23 | [0.06, 0.89] | 0.035 |
| Insurance type (Private) | b | 15.9095 | ~8.0e6 | [~0, ∞] | 0.987 |
| Insurance type (Public) | b | 16.2967 | ~1.1e7 | [~0, ∞] | 0.986 |
| Chronic ventilator use | c | 1.3934 | 4.03 | [2.23, 7.29] | <0.001 |
| Age at tracheostomy placement (years) | c | -0.1327 | 0.88 | [0.81, 0.96] | 0.002 |
| BPD diagnosis | c | -0.5782 | 0.56 | [0.30, 1.02] | 0.059 |
| HI-NI diagnosis | c | 0.5641 | 1.76 | [0.92, 3.36] | 0.087 |
| Male sex | c | 0.2710 | 1.31 | [0.78, 2.20] | 0.307 |
| Race (Other) | c | -0.3499 | 0.70 | [0.23, 2.12] | 0.547 |
| Race (White) | c | 0.0151 | 1.02 | [0.54, 1.93] | 0.964 |
| Non-Hispanic ethnicity | c | -1.2544 | 0.29 | [0.07, 1.13] | 0.072 |
| Insurance type (Private) | c | 14.5129 | ~2.0e6 | [~0, ∞] | 0.981 |
| Insurance type (Public) | c | 14.9402 | ~3.0e6 | [~0, ∞] | 0.981 |

## References

1. Russell CJ, Thurm C, Hall M, Simon TD, Neely MN, Berry JG. Risk factors for hospitalizations due to bacterial respiratory tract infections after tracheotomy. Pediatr Pulmonol. Jan 4 2018;doi:10.1002/ppul.23938.

2. Zhu H, Das P, Roberson DW, et al. Hospitalizations in children with preexisting tracheostomy: a national perspective. Laryngoscope. Feb 2015;125(2):462–8. doi:10.1002/lary.24797.

3. Watters KF. Tracheostomy in Infants and Children. Respir Care. 2017 Jun;62(6):799–825. doi: 10.4187/respcare.05366. PMID: 28546379.

4. Funamura JL, Yuen S, Kawai K, et al. Characterizing mortality in pediatric tracheostomy patients. Laryngoscope. Jul 2017;127(7):1701–1706. doi:10.1002/lary.26361.

5. Morrison JM, Hassan A, Kysh L, Dudas RA, Russell CJ. Diagnosis, management, and outcomes of pediatric tracheostomy-associated infections: A scoping review. Pediatric Pulmonology. May 2021.

6. Ishihara T, Tanaka H. Factors affecting tracheostomy in critically ill paediatric patients in Japan: a data-based analysis. BMC Pediatr. 2020 May 20;20(1):237. doi: 10.1186/s12887-020-02144-3. PMID: 32434537; PMCID: PMC7237622.

7. Powell J, Powell S, Mather MW, Beck L, Nelson A, Palmowski P, Porter A, Coxhead J, Hedley A, Scott J, Rostron AJ, Hellyer TP, Zaidi F, Davey T, Garnett JP, Agbeko R, Ward C, Stewart CJ, Taggart CC, Brodlie M, Simpson AJ. Tracheostomy in children is associated with neutrophilic airway inflammation. Thorax. 2023 Oct;78(10):1019–1027. doi: 10.1136/thorax-2022-219557. Epub 2023 Feb 20. PMID: 36808087; PMCID: PMC10511973.

8. Rusakow LS, Guarin M, Wegner CB, Rice TB, Mischler EH. Suspected respiratory tract infection in the tracheostomized child: the pediatric pulmonologist’s approach. Chest. Jun 1998;113(6):1549–54.

9. Cline JM, Woods CR, Ervin SE, Rubin BK, Kirse DJ. Surveillance tracheal aspirate cultures do not reliably predict bacteria cultured at the time of an acute respiratory infection in children with tracheostomy tubes. Chest. 2012 Mar;141(3):625–631. doi: 10.1378/chest.10-2539. Epub 2011 Mar 24. PMID: 21436240.

10. Steuart R, Ale GB, Woolums A, Xia N, Benscoter D, Russell CJ, Shah SS, Thomson J. Respiratory culture organism isolation and test characteristics in children with tracheostomies with and without acute respiratory infection. Pediatr Pulmonol. 2023 May;58(5):1481–1491. doi: 10.1002/ppul.26349. Epub 2023 Feb 15. PMID: 36751142.

11. Morar P, Singh V, Makura Z, Jones AS, Baines PB, Selby A, Sarginson R, Hughes J, van Saene R. Oropharyngeal carriage and lower airway colonisation/infection in 45 tracheotomised children. Thorax. 2002 Dec;57(12):1015–20. doi: 10.1136/thorax.57.12.1015. PMID: 12454294; PMCID: PMC1758797.

12. Warniment A, Steuart R, Rodean J, Hall M, Chinchilla S, Shah SS, Thomson J. Variation in Bacterial Respiratory Culture Results in Children With Neurologic Impairment. Hosp Pediatr. 2021 Nov;11(11):e326–e333. doi: 10.1542/hpeds.2020-005314. PMID: 34716209.

13. McCaleb R, Warren RH, Willis D, Maples HD, Bai S, O’Brien CE. Description of Respiratory Microbiology of Children With Long-Term Tracheostomies. Respir Care. 2016 Apr;61(4):447–52. doi: 10.4187/respcare.03518. Epub 2015 Dec 15. PMID: 26670471.

14. Wasserman MG, Graham RJ, Mansbach JM. Airway Bacterial Colonization, Biofilms and Blooms, and Acute Respiratory Infection. Pediatr Crit Care Med. 2022 Oct 1;23(10):e476–e482. doi: 10.1097/PCC.0000000000003017. Epub 2022 Jun 29. PMID: 35767569; PMCID: PMC9529803.

15. Zemanick ET, Emerson J, Thompson V, McNamara S, Morgan W, Gibson RL, Rosenfeld M; EPIC Study Group. Clinical outcomes after initial pseudomonas acquisition in cystic fibrosis. Pediatr Pulmonol. 2015 Jan;50(1):42–8. doi: 10.1002/ppul.23036. Epub 2014 Mar 18. PMID: 24644274.

16. Steuart R, Pan AY, Woolums A, et al. Respiratory culture growth and 3-year lung health outcomes in children with bronchopulmonary dysplasia and tracheostomies. Pediatr Pulmonol. 2024;59(2):300–313. doi:10.1002/ppul.26746.

17. Thomson JE, Feinstein JA, Hall M, Gay JC, Butts B, Berry JG. Identification of Children With High-Intensity Neurological Impairment. JAMA Pediatr. 2019 Oct 1;173(10):989–991. doi: 10.1001/jamapediatrics.2019.2672. PMID: 31424541; PMCID: PMC6704733.

18. Baron RM, Kenny DA. The moderator-mediator variable distinction in social psychological research: conceptual, strategic, and statistical considerations. J Pers Soc Psychol. 1986 Dec;51(6):1173–82. doi: 10.1037//0022-3514.51.6.1173. PMID: 3806354.

19. Russell CJ, Simon TD, Mamey MR, Newth CJL, Neely MN. Pseudomonas aeruginosa and post-tracheotomy bacterial respiratory tract infection readmissions. Pediatr Pulmonol. 2017 Sep;52(9):1212–1218. doi: 10.1002/ppul.23716. Epub 2017 Apr 25. PMID: 28440922; PMCID: PMC5561001.

20. Russell CJ, Simon TD, Neely MN. Development of Chronic Pseudomonas aeruginosa-Positive Respiratory Cultures in Children with Tracheostomy. Lung. 2019 Dec;197(6):811–817. doi: 10.1007/s00408-019-00285-6. Epub 2019 Oct 31. PMID: 31673781; PMCID: PMC6934374.

21. Jacobs DM, Ochs-Balcom HM, Noyes K, Zhao J, Leung WY, Pu CY, Murphy TF, Sethi S. Impact of Pseudomonas aeruginosa Isolation on Mortality and Outcomes in an Outpatient Chronic Obstructive Pulmonary Disease Cohort. Open Forum Infect Dis. 2020 Jan 4;7(1):ofz546. doi: 10.1093/ofid/ofz546. PMID: 31993457; PMCID: PMC6979313.

22. Rodrigo-Troyano A, Melo V, Marcos PJ, Laserna E, Peiro M, Suarez-Cuartin G, Perea L, Feliu A, Plaza V, Faverio P, Restrepo MI, Anzueto A, Sibila O. Pseudomonas aeruginosa in Chronic Obstructive Pulmonary Disease Patients with Frequent Hospitalized Exacerbations: A Prospective Multicentre Study. Respiration. 2018;96(5):417–424. doi: 10.1159/000490190. Epub 2018 Jul 24. PMID: 30041176.

23. Ashkenazi-Hoffnung L, Ari A, Bilavsky E, Scheuerman O, Amir J, Prais D. Pseudomonas aeruginosa identified as a key pathogen in hospitalised children with aspiration pneumonia and a high aspiration risk. Acta Paediatr. 2016 Dec;105(12):e588–e592. doi: 10.1111/apa.13523. Epub 2016 Aug 12. PMID: 27387674.

24. Chang I, Schibler A. Ventilator Associated Pneumonia in Children. Paediatr Respir Rev. 2016 Sep;20:10–16. doi: 10.1016/j.prrv.2015.09.005. Epub 2015 Sep 25. PMID: 26527358.

